# Insufficient serological evidence of the association between chronic kidney disease and leptospirosis in Badulla and Kandy districts, Sri Lanka

**DOI:** 10.1101/2023.05.31.23290784

**Authors:** Regina Amanda Fonseka, Pavani Senarathne, Devinda Shameera Muthusinghe, Nishantha Nanayakkara, Lishantha Gunaratne, Kumiko Yoshimatsu, Nobuo Koizumi, Chandika Damesh Gamage

## Abstract

**Objective:** Chronic kidney disease (CKD) and chronic kidney disease of uncertain etiology (CKDu) are chronic kidney diseases that pose a significant health burden in Sri Lanka. Leptospirosis is a bacterial zoonosis that primarily damages renal tissues by colonization of *Leptospira* spp. in the renal tubules and is a suspected etiological agent of CKDu. Since Sri Lanka is an endemic for leptospirosis and outbreaks of the disease have been reported, this study aimed to determine the association between leptospirosis and chronic kidney disease in two geographically distinct regions of Sri Lanka, Badulla (CKDu endemic) and Kandy (CKDu non-endemic) districts.

**Results:** Eighty-five patients with CKDu and 149 controls from Badulla and 49 patients with CKD and 135 controls from Kandy were serologically tested by microscopic agglutination test with a panel of 11 *Leptospira* serogroups. The seroprevalence rates for leptospirosis were 7.1% and 13.4% in the CKDu and control groups, respectively, in Badulla and 2.1% and 18.5% in the CKD and control groups, respectively, in Kandy. There were no statistically significant differences between demographic characteristics and leptospirosis seropositivity in the CKD and control groups in either Badulla or Kandy.

## Introduction

Chronic kidney disease (CKD) is a global public health problem primarily caused by due to diabetes and hypertension. CKD is defined as any condition that damages kidney tissue and results in a decrease in renal function with a glomerular filtration rate (GFR) of <60 mL/min/1.73 m^2^ for ≥ 3 months [1]. CKD that occurs in the absence of diabetes, hypertension, and other known risk factors is referred to as chronic kidney disease of uncertain etiology (CKDu) [2]. CKDu has been reported in Central America, Egypt, India, and Sri Lanka [3–6]. While several hypotheses are being explored to determine the causative agents/factors responsible for CKDu, one hypothesis that has been overlooked in Sri Lanka is an infectious etiology, such as hantavirus infection and leptospirosis [7].

Previous studies have implicated leptospirosis as a possible etiological agent or risk factor for CKDu [7–10]. Leptospirosis is one of the most prevalent bacterial zoonoses worldwide and is caused by spirochetes of the genus *Leptospira* [7]. The kidney is the primary target organ of *Leptospira* spp. If left untreated, the bacteria can colonize the renal tubules of the kidney tissue [10]. Renal colonization by *Leptospira* spp. can lead to interstitial tubular nephritis and acute kidney injury (AKI), which carry a risk for CKD [10]. Long-term mild subclinical AKI may lead to the development of CKD [11]. In 2015, an association between CKD and leptospirosis was first suggested in Taiwan: participants exposed to *Leptospira* spp. showed a 2.5% decrease in estimated GFR and a higher percentage of CKD compared to those not exposed to the bacteria [12]. In the same study, 88 individuals with high anti-leptospiral antibody titers were followed up for two years, and individuals with microscopic agglutination test (MAT) titers > 400 showed a positive correlation with the kidney injury marker KIM-1/Cr [12]. In 2018, Yang presented two hypothetical pathways by which leptospirosis could progress to CKD: acute *Leptospira* infection could progress to chronic leptospirosis if left untreated, leading to the development of CKD, while subclinical *Leptospira* infection could lead to chronic leptospirosis that may progress to CKD [10]. Heat stress and dehydration have been postulated to have synergistic effects on these pathways [10]. Furthermore, there are several similarities between leptospirosis kidney disease and CKDu, such as tubulointerstitial nephritis, interstitial fibrosis, non-proteinuria, proximal tubule dysfunction, and hypokalemia, and both diseases affect middle-aged men living in hot climates [10].

Although leptospirosis is an endemic and notifiable disease in Sri Lanka, it is still largely underdiagnosed and therefore most likely untreated, which can lead to a chronic state of infection without symptoms. The highest burden of CKDu occurs in the North Central Province (NCP), where leptospirosis outbreaks are also common [13, 14]. Since most of the CKDu affected population in the NCP are middle-aged men working in agriculture, it is possible that the renal tissue damage caused by *Leptospira* infection may progress to a chronic state due to incomplete recovery from mild AKI, recurrent exposure to *Leptospira* spp., and heat stress and dehydration. Therefore, this study investigated the involvement of leptospirosis as a causative agent or risk factor for CKD by comparing the seroprevalence of leptospirosis between chronic kidney disease patients and healthy controls in a CKDu endemic district, Badulla, and a CKDu non-endemic district, Kandy, Sri Lanka.

## Methods

### Study design

This is a hospital and community-based cross-sectional study with unmatched cases and controls. The study areas were Mahiyangana Divisional Secretariat, Badulla district, an endemic region of CKDu, and Yatinuwara Divisional Secretariat, Kandy district, a non-endemic region of CKDu, Sri Lanka.

### Sample collection

Sample collection was conducted between January 2017 and February 2018. Clinically diagnosed CKDu patients aged ≥18 years residing in Badulla (n=85) and CKD patients aged ≥18 years in Kandy (n=49) districts and attending the renal clinics of the Nephrology and Transplantation Unit, Girandurukotte District Hospital (Badulla) and Teaching Hospital Kandy (Kandy), respectively, were recruited as cases for the study. Individuals with a history of alcoholism were excluded from the study. For the control groups, the Medical Office of Health areas within the Badulla and Kandy districts were randomly selected. Individuals aged ≥18 years, residing in Badulla (n=149) and Kandy (n=135) districts, with no clinically diagnosed renal disease and normal serum creatinine levels (0.5– 1.2 mg/dL) volunteered to participate in this study. Individuals with a history of either alcoholism or kidney disease were excluded from the study.

Five milliliters of blood were collected in sterile tubes without anticoagulant. The tubes were left undisturbed at room temperature for 30 min and centrifuged at 2000×*g* for serum separation. Serum separation was performed at the sample collection site and immediately transported in insulated coolers to the Department of Microbiology, Faculty of Medicine, University of Peradeniya, and serum samples were stored at −20°C prior to analysis.

At the time of sample collection, demographic information was collected using a structured questionnaire. The questionnaire covered the following information: basic demographic data (age and sex), family and past medical history, occupational information, agricultural activity, and exposure to rodents.

### Detection of anti-leptospiral antibodies by MAT

The MAT panel antigens used in this study were obtained from the Veterinary Research Institute and contained 12 serovars belonging to 11 serogroups: serogroup Autumnalis (serovar Autumnalis), Bataviae (Bataviae), Canicola (Canicola), Grippotyphosa (Grippotyphosa), Hebdomadis (Hebdomadis), Javanica (Javanica), Panama (Panama), Sejroe (Hardjo and Wolffi), Semaranga (Patoc), Shermani (Shermani), and Tarassovi (Tarassovi). The strains were maintained in a liquid Ellinghausen McCullough Johnson and Harris medium supplemented with *Leptospira* enrichment (BD Difco, USA) and 5-flurouracil (final concentration of 200 µg/mL). Four- to seven-day-old cultures with growth equivalent to 0.5 McFarland (approximately 1-2×10^8^ organisms/mL) were used for MAT as previously described [15, 16]. A MAT titer of ≥1:400 was considered as positive for acute leptospirosis, as a titer of ≥1:320, is considered clinically significant in Sri Lanka [17].

### Statistical analysis

All statistical analyses were performed using the Statistical Package for the Social Sciences version 25 (IBM, USA) and the online tool MedCalc (https://www.medcalc.org/). Exposure to *Leptospira* spp. and possible risk factors were evaluated by 2×2 Yate’s corrected chi-square test or Fisher’s exact test. A P value less than 0.05 was considered statistically significant. The odds ratio and associated 95% confidence interval (CI) were calculated in the exposed group when compared with the non-exposed group for *Leptospira* spp.

## Results

### Leptospirosis seroprevalence in Badulla and Kandy

The seroprevalence rates in the CKDu and control groups in Badulla were 7.1% and 13.4%, respectively (Table 1). There was no statistically significant difference between the disease and control groups (p=0.14). Seroprevalences of 2.0% and 18.5% for leptospirosis were observed in the CKD and control groups, respectively, in Kandy (Table 1). The seroprevalence in the CKD group was significantly lower than that in the control group (p=0.02).

**Table 1.** Leptospirosis seropositivity between CKD patients and healthy individuals in Badulla and Kandy.

| District | Group | No. of tested | No. of MAT positives (%) | Odds Ratio (95% CI) | P value |
| --- | --- | --- | --- | --- | --- |
| Badulla | CKDu patients | 85 | 6 (7.1) | 0.49 (0.19–1.27) | 0.14 |
|  | Control | 149 | 20 (13.4) |  |  |
| Kandy | CKD patients | 49 | 1 (2.0) | 0.09 (0.01–0.70) | 0.02 |
|  | Control | 135 | 25 (18.5) |  |  |

### Comparison of demographic characteristics and seropositivity in Badulla and Kandy

There were no statistically significant differences in demographic characteristics and leptospirosis seropositivity between the CKD and control groups in Badulla and Kandy (Table 2).

**Table 2.**
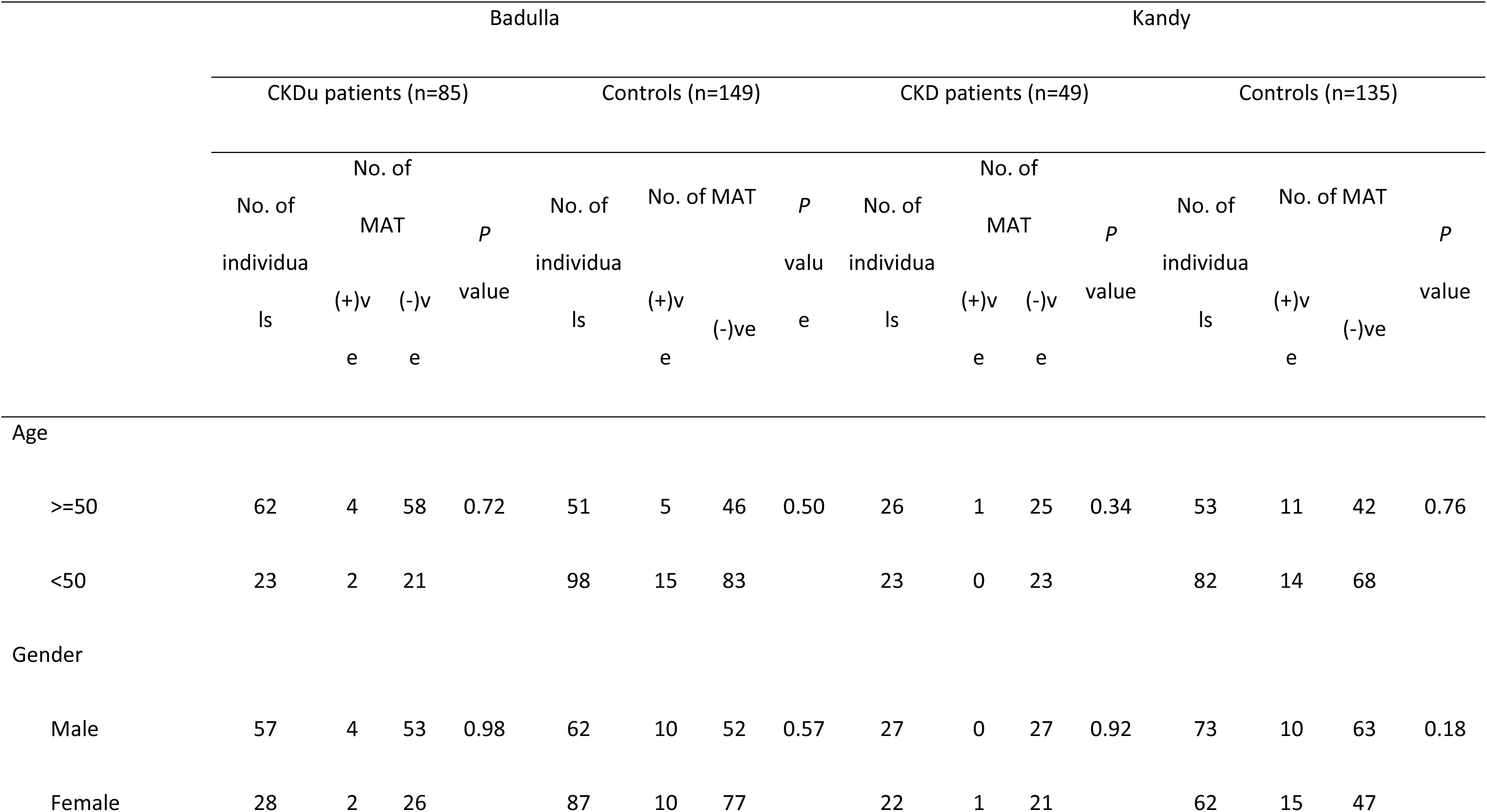
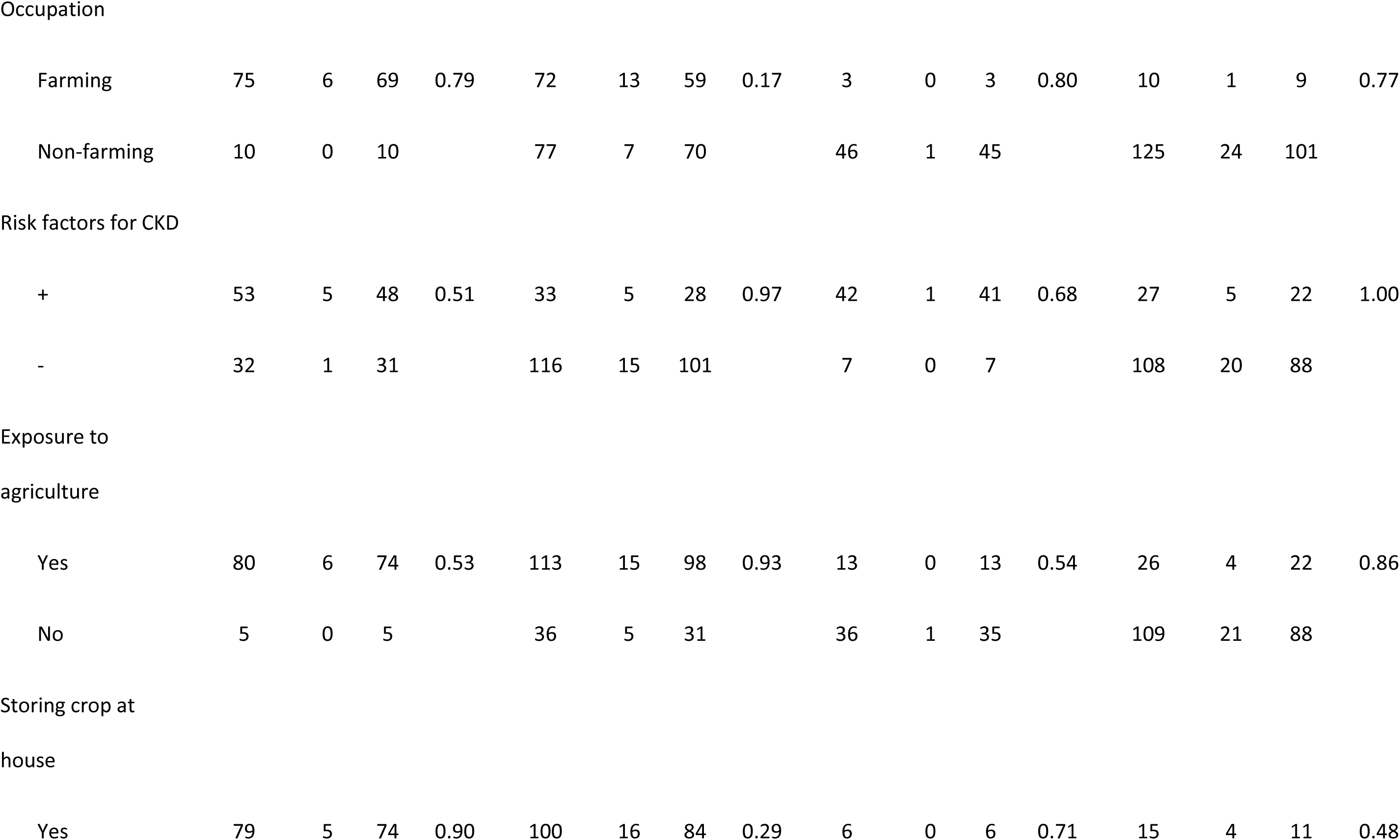
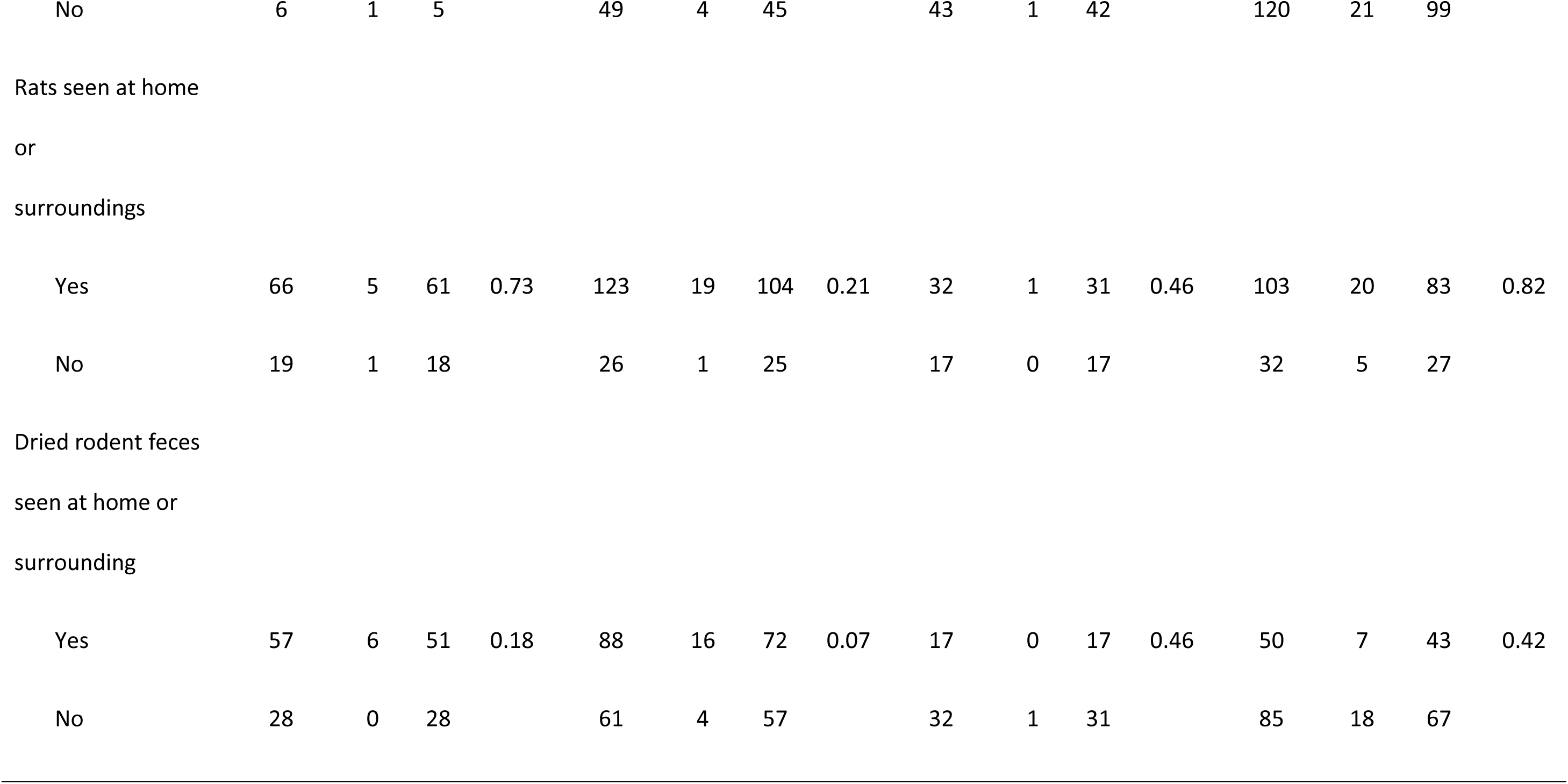
Comparison of demographic characteristics and seropositivity in CKD patients and controls in Badulla and Kandy.

### Infecting *Leptospira* serogroups in Badulla and Kandy

Serogroup Sejroe was the predominant infecting serogroup in both Badulla and Kandy (Table 3). Although a MAT titer ≥1:400 was considered positive for leptospiral antibody indicating acute infection in this study, individuals with lower antibody titers were also observed (Supplementary Table 1).

**Table 3.**
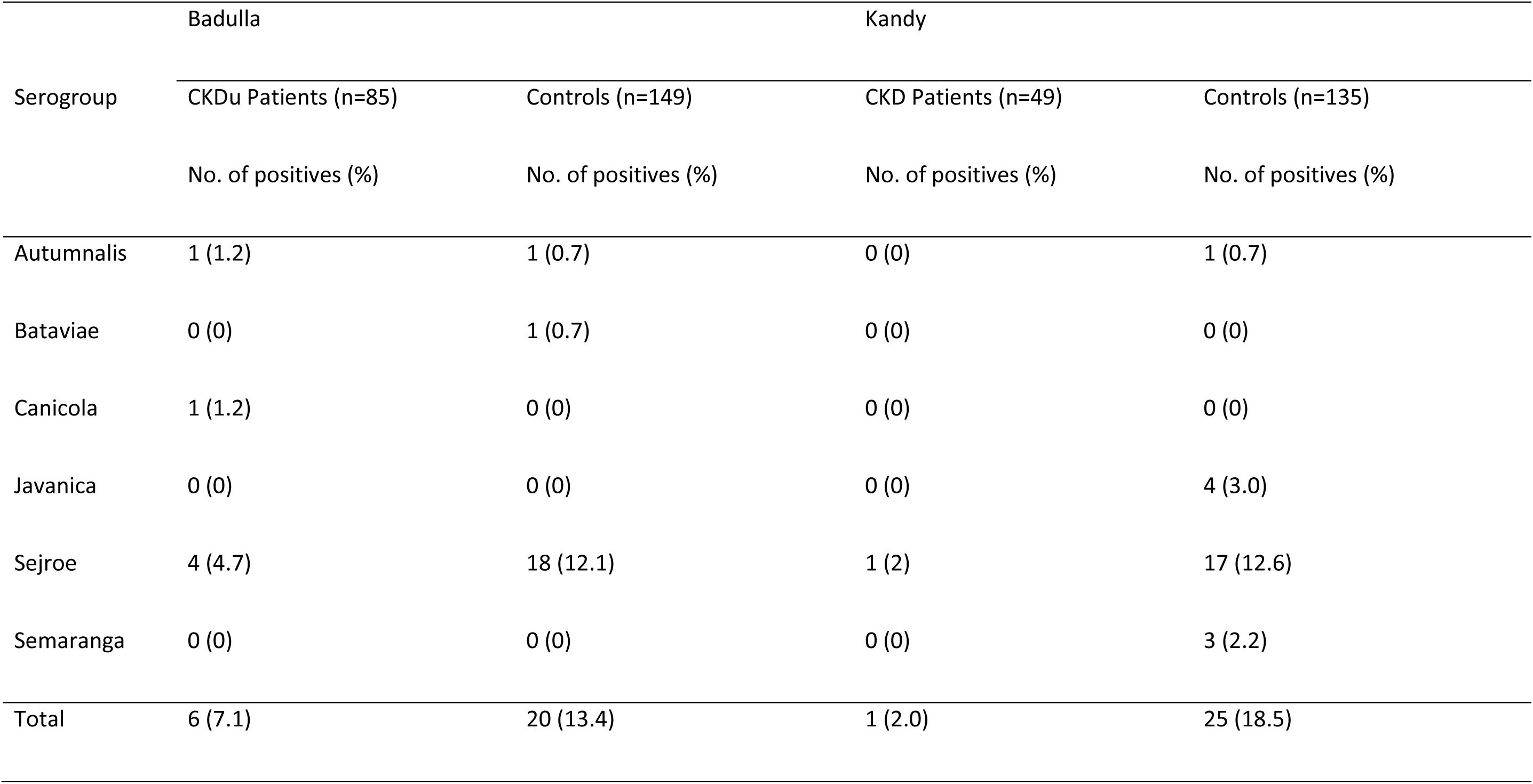
Results of MAT in Badulla and Kandy.

## Discussion

This study suggests that there is no association between CKD and leptospirosis seroprevalence. This finding is consistent with that of a recent study conducted in the NCP of Sri Lanka in 2020, where the difference in leptospirosis seroprevalence between the CKDu and control groups was not statistically significant [18]. On the other hand, these results contradict the findings of a study conducted in Taiwan, which showed an association between *Leptospira* seroprevalence and CKD and lower eGFR [12]. The cause of CKDu may vary regionally and be geographically specific. Epidemics of kidney disease of unknown etiology have previously been reported: Itai-itai disease and Balkan endemic nephropathy (BEN) are nephropathic epidemics whose etiology was identified much later. Itai-itai disease, which occurred in Japan in 1912, was found to be caused by cadmium poisoning in 1968, while BEN, which occurred in the 1950s, was found to be caused by aristocholic acid in 1993 [19].

This study showed that the predominant infecting *Leptospira* serogroup was Sejroe, of which serovar Hardjo was the most reactive serovar in both Badulla and Kandy (Table 3 and Supplementary Table 1). Serovar Hardjo is known to be maintained by cattle and the transmission of this strain to humans often occurs through contact with cattle [20]. The high seroprevalence for serogroup Sejroe (serovar Hardjo) can be explained by the exposure of the population to agricultural activities with direct/indirect contact with cattle in Badulla (Table 2). However, a high seroprevalence for serogroup Sejroe (serovar Hardjo) was also observed in the Kandy population, although most of them were not involved in agriculture (Table 2). Hardjo infections in dogs and cats have been reported previously in Brazil, Croatia, Italy, Scotland, and the USA [21–27]. In addition, a previous serological study of companion dogs in Kandy district showed that Sejroe was the most prevalent serogroup [16].

### Limitations

The CKD cohort in this study consisted of CKD and CKDu patients, whereas it would have been ideal to recruit only CKDu patients. Differentiation between CKDu and CKD patients is possible only by renal biopsy, an invasive procedure that is rarely performed in Sri Lanka. The antigen panel used for MAT was not composed of locally isolated strains. It has been shown that renal colonization with *Leptospira* spp. can occur asymptomatically and sometimes in the absence of serological evidence [12, 28, 29]. To conclude that leptospirosis is not associated with CKD in Sri Lanka, renal colonization needs to be investigated.

## Data Availability

Data has been made available as Supplementary Materials

## List of abbreviations

AKI: acute kidney injury
CI: confidence interval
CKD: chronic kidney disease
CKDu: chronic kidney disease of uncertain etiology
GFR: glomerular filtration rate
MAT: microscopic agglutination test
NCP: North Central Province

## Declarations

### Ethics approval and consent to participate

Ethical approval for this study was obtained from the institutional ethical review committee of the Faculty of Medicine, University of Peradeniya (2016/EC/64). The collection of blood samples and demographic data was approved by the Regional Director of Health Services, Kandy, and informed consent was obtained from all study participants via signature or thumbprints.

### Consent for publication

Not applicable

### Availability of data and materials

All data generated or analyzed during this study are included in this published article and its supplementary information file.

### Authors’ contributions

Conceptualization: CDG; Investigation: RAF, PS; Formal analysis: RAF, CDG; Data curation: RAF, PS; Project administration: RAF, PS, NN, LG; Methodology: NK, CDG; Resources: KY, NK, CDG; Supervision: NK, CDG; Funding acquisition: NK, CDG; Writing - Original Draft: RAF; Writing - Review & Editing: RAF, PS, DSM, NN, LG, YK, NK, CDG

## Acknowledgments

We thank Athula Kumara and all the staff members of the Department of Microbiology, Faculty of Medicine, and Nishanthi Weerakoon and all the staff members of the Microbiology Division of the Veterinary Research Institute, for their technical support. We also thank Yomani Sarathkumara for collecting samples and data.

## References

1. Levey AS, Eckardt KU, Tsukamoto Y, Levin A, Coresh J, Rossert J, et al. Definition and classification of chronic kidney disease: A position statement from Kidney Disease: Improving Global Outcomes (KDIGO). Kidney Int. 2005;67:2089–100.

2. Weaver VM, Fadrowski JJ, Jaar BG. Global dimensions of chronic kidney disease of unknown etiology (CKDu): a modern era environmental and/or occupational nephropathy? BMC Nephrol. 2015;16:145.

3. González-Quiroz M, Pearce N, Caplin B, Nitsch D. What do epidemiological studies tell us about chronic kidney disease of undetermined cause in Meso-America? A systematic review and meta-analysis. Clin Kidney J. 2018;11:496–506.

4. O’Callaghan-Gordo C, Shivashankar R, Anand S, Ghosh S, Glaser J, Gupta R, et al. Prevalence of and risk factors for chronic kidney disease of unknown aetiology in India: Secondary data analysis of three population-based cross-sectional studies. BMJ Open. 2019;9: e023353.

5. Ruwanpathirana T, Senanayake S, Gunawardana N, Munasinghe A, Ginige S, Gamage D, et al. Prevalence and risk factors for impaired kidney function in the district of Anuradhapura, Sri Lanka: a cross-sectional population-representative survey in those at risk of chronic kidney disease of unknown aetiology. BMC Public Health. 2019;19:763.

6. El Minshawy O. End-stage renal disease in the El-Minia Governorate, upper Egypt: an epidemiological study. Saudi J Kidney Dis Transpl. 2011;22:1048–54.

7. Gamage CD, Sarathkumara YD. Chronic kidney disease of uncertain etiology in Sri Lanka: Are leptospirosis and Hantaviral infection likely causes? Med Hypotheses. 2016;91:16–9.

8. Carrillo-Larco RM, Altez-Fernandez C, Acevedo-Rodriguez JG, Ortiz-Acha K, Ugarte-Gil C. Leptospirosis as a risk factor for chronic kidney disease: A systematic review of observational studies. PLoS Negl Trop Dis. 2019;13: e0007458.

9. Correa-Rotter R, Wesseling C, Johnson RJ. CKD of unknown origin in Central America: the case for a Mesoamerican nephropathy. Am J Kidney Dis. 2014;63:506–20.

10. Yang CW. Leptospirosis renal disease: emerging culprit of chronic kidney disease unknown etiology. Nephron. 2018;138:129–36.

11. Kupferman J, Ramírez-Rubio O, Amador JJ, López-Pilarte D, Wilker EH, Laws RL, et al. Acute kidney injury in sugarcane workers at risk for Mesoamerican nephropathy. Am J Kidney Dis. 2018;72:475–82.

12. Yang HY, Hung CC, Liu SH, Guo YG, Chen YC, Ko YC, et al. Overlooked risk for chronic kidney disease after leptospiral infection: a population-based survey and epidemiological cohort evidence. PLoS Negl Trop Dis. 2015;9: e0004105.

13. Wanigasuriya K. Update on uncertain etiology of chronic kidney disease in Sri Lanka’s north-central dry zone. MEDICC Rev. 2014;16:61–5.

14. Agampodi SB, Dahanayaka NJ, Bandaranayaka AK, Perera M, Priyankara S, Weerawansa P, et al. Regional differences of leptospirosis in Sri Lanka: observations from a flood-associated outbreak in 2011. PLoS Negl Trop Dis. 2014;8: e2626.

15. Brandão AP, Camargo ED, da Silva ED, Silva MV, Abrão. Macroscopic agglutination test for rapid diagnosis of human leptospirosis. J Clin Microbiol. 1998;36; 3138–42.

16. Athapattu T, Fernando R, Abayawansha R, Fernando P, Fuward M, Samarakoon N, et al. Carrier status of *Leptospira* spp. in healthy companion dogs in Sri Lanka. Vector Borne Zoonotic Dis. 2022;22:93–100.

17. Epidemiology Unit E, Ministry of Health N and IMSL. National guidelines on management of leptospirosis. Epidemiology Unit, Epidemiology Ministry of Health, Nutrition and Indigenous Medicine Sri Lanka. 2016.

18. Sunil-Chandra NP, Jayaweera JAAS, Kumbukgolla W, Jayasundara MVML. Association of hantavirus infections and leptospirosis with the occurrence of chronic kidney disease of uncertain etiology in the North Central Province of Sri Lanka: a prospective study with patients and healthy persons. Front Cell Infect Microbiol. 2020;10: 556737.

19. Gifford FJ, Gifford RM, Eddleston M, Dhaun N. Endemic nephropathy around the world. Kidney Int Rep. 2017;2:282–92.

20. White FH, Sutherland GE, Raynor LE, Cottrell CR, Sulzer KR. *Leptospira interrogans serovars hardjo* and *pomona*: causes of infections in dairy cows and humans in Florida. Public Health Rep. 1981;96:250–4.

21. Adin CA, Cowgill LD. Treatment and outcome of dogs with leptospirosis: 38 cases (1990-1098). J Am Vet Med Assoc. 2000;216:371–5.

22. Agunloye CA, Nash AS. Investigation of possible leptospiral infection in cats in Scotland. J Small Anim Pract. 1996;37:126–9.

23. Kikuti M, Langoni H, Nobrega DN, Corrêa APFL, Ullmann LS. Occurrence and risk factors associated with canine leptospirosis. J Venom Anim Toxins Incl Trop Dis. 2012;18:124–7.

24. Majetić ZŠ, Habuš J, Milas Z, Perko VM, Starešina V, Turk N. Serological survey of canine leptospirosis in Croatia - the changing epizootiology of the disease. Vet Arh. 2012;82:183–91.

25. Piredda I, Ponti MN, Piras A, Palmas B, Pintore P, Pedditzi A, et al. New insights on *Leptospira* infections in a canine population from north Sardinia, Italy: a sero-epidemiological study. Biology. 2021;10:507.

26. Rubanick J V., Fries RC, Waugh CE, Pashmakova MB. Severe hyperkalemia presenting with wide-complex tachycardia in a puppy with acute kidney injury secondary to leptospirosis. J Vet Emerg Crit Care. 2016;26:858–63.

27. Shropshire SB, Veir JK, Morris AK, Lappin MR. Evaluation of the *Leptospira* species microscopic agglutination test in experimentally vaccinated cats and *Leptospira* species seropositivity in aged azotemic client-owned cats. J Feline Med Surg. 2016;18:768–72.

28. Ganoza CA, Matthias MA, Saito M, Cespedes M, Gotuzzo E, Vinetz JM. Asymptomatic renal colonization of humans in the Peruvian Amazon by *Leptospira*. PLoS Negl Trop Dis. 2010;4:e612.

29. Sivasankari K, Shanmughapriya S, Natarajaseenivasan K. Leptospiral renal colonization status in asymptomatic rural population of Tiruchirapalli district, Tamilnadu, India. Pathog Glob Health. 2016;110:209–15.

